# Acceptability of pre-exposure prophylaxis and associated factors among HIV-negative young men in Kagwara fishing community- Serere district: a cross-sectional study

**DOI:** 10.1101/2025.01.07.25320147

**Authors:** Alex Omoding, Ronald Opito, Paul Oboth, Francis Okello, Joseph KB Matovu

## Abstract

**Background:** Despite the potential efficacy of Pre-Exposure Prophylaxis (PrEP) in reducing HIV risk among at-risk populations, PrEP acceptability remains strikingly varied across populations. We assessed PrEP acceptability and associated factors among at-risk HIV-negative young men in Kagwara fishing community, Serere district.

**Methods:** A cross-sectional quantitative study design was used. Data were collected among 409 at-risk HIV-negative young men aged 15-24 years living in Kagwara fishing community, between August and October 2023. Quantitative data were collected on socio-demographic characteristics, sexual-risk behaviors, knowledge, attitudes, and practices regarding PrEP acceptability. PrEP acceptability was defined as the proportion of young men accepting to use PrEP out of those interviewed, based on six constructs adopted from acceptability framework. Data analyzed using Stata version 15.0 statistical software. Summary statistics computed and presented as tables, frequencies and proportions. Bivariate analysis was conducted using penalized logistic regression to identify independent factors associated with PrEP acceptability. All factors that had p<0.10 at the bivariate analysis and suspected confounders were entered into the final logistic regression model. All factors with p<0.05 were significantly associated with the primary outcome.

**Results:** Of 409 respondents, average age was 21.8(SD=1.9) years. Majority, (97.8%, n=393) had unprotected penetrative sex while 84.6% (n=346) did not know HIV status of their partners. PrEP acceptability was high at 93.6% (n=383) as majority of the participants accepted to use PrEP based on the six constructs of acceptability. At multivariable level, the factors associated with PrEP acceptability were; perceived risk of getting HIV infection, adjusted odds ratio (aOR)(95%CI)=4.23(1.05, 17.04), knowing the partner’s HIV status, aOR (95%CI) = 0.25 (0.07, 0.88), feeling embarrassed to ask for PrEP from the facility, aOR (95%CI) = 0.12 (0.04, 0.39), and having concern of stigma associated with use of PrEP, aOR = 0.13 (95% CI, 0.04-0.41).

**Conclusion:** We found a high level of PrEP acceptability among at-risk HIV-negative young men in Kagwara fishing community. Improving access to PrEP services among at-risk young men in the fishing communities may increase PrEP uptake in this population and across similar settings.

## Introduction

Despite the availability of recognized and recommended Pre-Exposure Prophylaxis (PrEP) for HIV prevention among at-risk populations, new HIV infections have persisted and are highest among adolescents and young people [1]. Globally, 1.3 million people were newly infected with HIV in 2022 and infections occurring among young people aged 15-24 years contributed 20% [2]. In Uganda, an average of 52,000 people become infected with HIV every year, with 36% of these new infections occurring among adolescents and young people [3]. The HIV prevalence in Uganda has a geographical heterogeneity with fishing communities listed among those with high prevalence [4]. The HIV prevalence among young fisher-folks is at least twice as high as among young people in the general population [3]. This could be attributed to the multiple co-occurring transitions during adolescence such as increased autonomy, decreased adult supervision, identity formation, peer influence, substance abuse (alcohol and other illicit drugs), emotional and social transition, potentially leading to early sexual debut and health risk behaviors that may lead to HIV infections [5–7]. In addition, fishing communities have increased susceptibility to HIV due to complex interacting factors like high mobility, poor access to healthcare including HIV prevention and care services, low perception of HIV risk, and poor access to HIV information [8]. Also, fishing communities are known for having sexually active persons who engage in risky sexual intercourse and high populations who engage in transactional sex [9–11], to which some young people get inclined [12,13].

Uptake of PrEP among at-risk people in the fishing communities remains largely undocumented, yet its success as an intervention greatly depends on its acceptability [14,15]. According to studies across several populations, PrEP acceptability has varied from as low as 1% among the young people in South Africa [16] to as high as above 90% among the female sex workers in Zambia [17]. Most PrEP implementation efforts have focused primarily on reaching at HIV high-risk populations, especially female sex workers, men who have sex with men, HIV discordant couples, adolescent girls and young women because they account for the majority of new HIV infections [17–21]. Subsequent studies on PrEP use in settings providing standard of care have also focused on the same populations leaving out young men who are also sexually active and are sexual counterparts of women who are at high-risk of HIV infection. Therefore, this study aimed at assessing acceptability of pre-exposure prophylaxis and associated factors among at-risk HIV-negative young men in Kagwara fishing community-Serere district to provide supportive evidence towards provision and scale up of PrEP use.

## Methods and Materials

### Study design

This study adopted a cross-sectional design, and a random sampling technique was used in which a representative population of at-risk HIV-negative young men in the fishing community of Kagwara was studied at one instance each. The design was chosen because it incorporated the use of questionnaire as the data capture tool, with the implication that it was possible to collect quantifiable data that was required to answer the research questions. In addition, collection of not only descriptive data that could be analyzed but also the possibility of analyzing relationships between any descriptive variables of interest [22,23]. In other words, the design allowed for the analysis of all possible factors associated with PrEP acceptability among the at-risk HIV negative young men.

## Study setting

This study was conducted in Kagwara fishing community; Serere district. Kagwara fishing community was chosen as the study site because just like other fishing communities; HIV incidence is high [24] and it is being targeted for PrEP provision yet the targeted persons therein have largely demonstrated hesitance towards HIV prevention methods for reasons not yet established by the ministry of health or its partners. There were certainly several associated factors to that behavior that needed to be assessed whether PrEP would be accepted before it is introduced. Kagwara fishing community is one of the fishing communities in Lake Kyoga found in Eastern part of Uganda. It is being served by Kagwara HC III, one of the health facilities in Kadungulu sub-county.

## Study population

The study population was young men who were sexually active, having concurrent multiple (two or more) sexual partners, young men reporting no or inconsistent condom use with women of unknown HIV status or HIV positive women and young men with known HIV-negative test result. Young men who were medically compromised to provide informed consent and young men who did not identify with their birth gender were excluded from the study.

## Study variables

### Dependent variable

The dependent variable (primary outcome) for this study was acceptability of HIV PrEP which was defined as proportion of at-risk HIV-negative young men that would express intention to use PrEP for HIV prevention purpose if made available to them at no cost. PrEP acceptability was derived as a composite variable from six out of seven theoretical constructs of acceptability framework [TFA] [25]. These included: affective attitude, burden, perceived effectiveness, intervention coherence, self-efficacy and opportunity cost. Although the acceptability framework includes seven theoretical constructs, we used six constructs because the seventh construct (Ethicality) had an ambiguous question which was not straight-forward and therefore confused the participants. Each theoretical construct had one question with five responses on a liked scale. The five responses were collapsed into two (2), that is, positive, scored one (1) and negative, scored as zero (0). This helped in the final computations, and it avoided scores cancelling the result. The sum of the six responses per participant was computed and ranged from zero (0) to six (6). Participants with scores of 4-6 were considered as having accepted PrEP while those with scores of 0-3 were considered as having not accepted PrEP.

### Independent variables

The independent variables in this study were factors associated with PrEP acceptability that included the sociodemographic characteristics such as age, marital status, education level, religion, behavioral characteristics such as having multiple sexual partners, knowing the partners’ HIV status, use of drugs of abuse and other medical related conditions such as having ever been diagnosed with sexually transmitted infection (STI).

## Data collection procedures and methods

The study participants were recruited and data collected between August-October 2023. An introductory letter was obtained from Busitema University and approval sought from the district health officer of Serere district. The approved introductory letter was then used for introducing research team to the management of the fishing community. The governor is the overall leader of the fishing community who works in conjunction with recognized government arms like local councils, police and local government officials like parish chief and sub-county chief. The young men were then accessed through these leaders of the fishing community. Their age eligibility (15-24 years) to participate in the study was verified using the documents obtained at the landing site from the governor but also others who mentioned verbally their years were considered and more so those who corroborated their years of birth with special events that had happened in the specified period of 15-24 years (1994-2008) and this helped in calculating the actual years.

On daily basis working with community leaders’ young men found doing activities of the day would be approached and the purpose of the visit explained. Those who accepted to participate in the study would randomly be selected and the subsequent procedures would then be conducted that included further screening for study eligibility, offering HIV counselling and testing and those who turned negative would then be interviewed following research questionnaire that had been configured in KOBO tool.

## Data analysis

Data were extracted from the KOBO tool and exported to STATA (version 15.0) statistical software for analysis. Data were analyzed at 3 different levels. Descriptive statistics such as mean, frequencies, standard deviations and range were done and presented as tables, frequencies and proportions. Bivariate analysis was conducted using penalized logistic regression to compare independent predictors of PrEP acceptability with the outcome of interest. The final multivariable analysis involved using penalized logistic regression to determine the factors associated with acceptability of PrEP among this population. These were reported as adjusted odds ratios (aOR) and factors whose confidence intervals of the OR did not include a null (1.0) were considered statistically significant.

## Results

### Socio-demographic characteristics of the respondents

Table 1 shows the socio-demographic characteristics of the study population. Overall, 409 respondents participated in the study. The average age of the respondents was 21.8 years (SD: ±1.9). Most (87.8%, n=359) of the respondents were aged 20-24 years and 12.2% (n= 50) were aged 15-19 years. About 61.9% (n=253) of the respondents had primary education as their highest level of education reached while 21.5% (n=88) had attended secondary education and 1% (n=4) had tertiary education and 15.6% (n= 64) of the respondents had no formal education. Slightly above three quarters (79%, n=323) were fishermen and more than half (68.4%, n=280) were married.

**Table 1.** Socio-demographic characteristics of the respondents.

| Variables | Frequency<br>(N=409) | Proportion<br>(%) |
| --- | --- | --- |
| Mean age/years | 21.8(SD=1.9) |  |
| **Age group (Complete years)** |  |  |
| 15-19 years | 50 | 12.2% |
| 20-24 years | 359 | 87.8% |
| **Education Level** |  |  |
| None | 64 | 15.6% |
| Primary | 253 | 61.9% |
| Secondary | 88 | 21.5% |
| Tertiary | 4 | 1.0% |
| **Occupation** |  |  |
| Business | 6 | 1.5% |
| Fishing | 323 | 79.0% |
| Peasant | 45 | 11.0% |
| Salaried | 2 | 0.5% |
| Self-employed | 2 | 0.5% |
| Student | 31 | 7.6% |
| **Marital Status** |  |  |
| Single | 129 | 31.6% |
| Married | 280 | 68.4% |

### Behavioral characteristics of the respondents

Table 2 shows the behavioral characteristics of the respondents. More than half (59.2% n=242) of the respondents had multiple sexual partners. Most (97.8%, n=393) of the respondents had un-protected sexual intercourse within six months of the study. About 84.6% (n=346) did not know their partner(s) HIV status. More than half (52.6%, n=215) used drugs of abuse and 61.1% (n=250) of the respondents had never heard about PrEP. All (100%, n= 409) of the respondents were aware of the availability of HIV services at the health facilities. Most (91.9%, n=376) of the respondents would not feel embarrassed to ask for PrEP from health facilities if made available freely. More than three quarters (78.5%, n=321) of the respondents would take PrEP because of perceived risk of HIV. About 63.1% (n=258) of the respondents would take PrEP because of having multiple sexual partners. Three quarters (76.5% n=313) of the respondents would take PrEP because of having sex without condoms. About 78.5% (n=321) of the respondents would take PrEP because of having sexual partners of unknown HIV status and most (95.6%, n= 391) of the respondents would take PrEP because of history of sexually transmitted disease.

**Table 2.** Behavioral characteristics of the respondents.

| Variable | Frequency | Percentage |
| --- | --- | --- |
| **Number of Sexual Partners** |  |  |
| One | 167 | 40.8% |
| More than one | 242 | 59.2% |
| **Was penetrative sex protected (N=404)** |  |  |
| No | 393 | 97.8% |
| Yes | 13 | 3.2% |
| **Know partner HIV status** |  |  |
| No | 346 | 84.6% |
| Yes | 63 | 15.4% |
| **Used addictive drugs in the last 6 months** |  |  |
| No | 215 | 52.6% |
| Yes | 194 | 47.4% |
| **Ever heard about PrEP** |  |  |
| No | 250 | 61.1% |
| Yes | 159 | 38.9% |
| **HIV services offered with other services at the facility** |  |  |
| No | 0 | 0.0% |
| Yes | 409 | 100.0% |
| **Feeling embarrassed to ask for PrEP at facility** |  |  |
| No | 376 | 91.9% |
| Yes | 33 | 8.1% |
| **Perceived risk of HIV infection motivates me to use PrEP** |  |  |
| No | 88 | 21.5% |
| Yes | 321 | 78.5% |
| **Having multiple sexual partners motivates me to use PrEP** |  |  |
| Yes | 258 | 63.1% |
| No | 151 | 36.9% |
| **Having sex without condoms motivates me to use PrEP** |  |  |
| Yes | 313 | 76.5% |
| No | 96 | 23.5% |
| **Having sexual partners of unknow HIV status motivates me to use PrEP** |  |  |
| Yes | 321 | 78.5% |
| No | 88 | 21.5% |
| **History of sexually transmitted disease** |  |  |
| Yes | 391 | 95.6% |
| No | 18 | 4.4% |

### Concerns about HIV PrEP

Table 3 shows concerns about HIV PrEP highlighted by the at-risk HIV negative young men in Kagwara fishing community. Almost three quarters (72.9%, n=298) of the respondents had concerns about PrEP. More than half (55.7%, n=166) of the respondents with concerns pointed out less effectiveness of below one hundred percent. About 58.1% (n=173) of respondents mentioned side effects. More than quarter (29.5%, n=88) of the respondents pointed out costs associated with PrEP. About 39.9% (n=119) of the respondents with concerns raised a concern of pill burden. Almost a third (31.9%, n=95) of the respondents pointed out stigma. About 12.4% (n= 37) of respondents were not comfortable of using PrEP while at the same time using condoms and a few 1.3% (n=4) of the respondents with concern pointed out the health workers negative attitude as some of the concerns about HIV PrEP.

**Table 3.** Concerns about HIV PrEP.

| Variable | Frequency | Percentage |
| --- | --- | --- |
| **Having any PrEP concerns** |  |  |
| No | 111 | 27.1% |
| Yes | 298 | 72.9% |
| **Effectiveness of less than 100% (N=298)** |  |  |
| No | 132 | 44.3% |
| Yes | 166 | 55.7% |
| **Side effects concern (N=298)** |  |  |
| No | 125 | 41.9% |
| Yes | 173 | 58.1% |
| **Cost concerns (N=298)** |  |  |
| No | 210 | 70.5% |
| Yes | 88 | 29.5% |
| **Pill burden concern (N=298)** |  |  |
| No | 179 | 60.1% |
| Yes | 119 | 39.9% |
| **Stigma concern (N=298)** |  |  |
| No | 203 | 68.1% |
| Yes | 95 | 31.9% |
| **Still must use condom concern (N=298)** |  |  |
| No | 261 | 87.6% |
| Yes | 37 | 12.4% |
| **Health workers negative receptivity concern (N=298)** |  |  |
| No | 294 | 98.7% |
| Yes | 4 | 1.3% |

*Key: The numbers in this table are varying because the first question was asked to all* *respondents, and the subsequent questions were asked only respondents who had concerns* *regarding PrEP*

## PrEP acceptability based on the constructs of theoretical framework of acceptability

Table 4 shows PrEP Acceptability based on 6 out of 7 constructs of the theoretical framework of acceptability (TFA). Overall, 93.6% (n=383) of the at-risk HIV negative young men in Kagwara fishing community accepted PrEP for HIV prevention purpose. Most (91.4%, n=373) of the respondents had positive attitude towards PrEP and were comfortable. Three quarters (76.5%, n=313) of the respondents felt there was no burden associated with PrEP and strongly mentioned that it will take little or no effort to use PrEP. About 82.4% (n=337) of the respondents had a good perception of PrEP effectiveness and pointed that PrEP is likely to reduce their chance of getting infected with HIV. Most (92.9%, n=380) of the respondents agreed that it was clear to them how using PrEP reduces chances of getting infected with HIV (intervention coherence). About 89.5% (n=366) of the respondents were confident of being able to use PrEP (self-efficacy) and most 90.9% (n=372) of the respondents disagreed that using PrEP would interfere with their other priorities (opportunity costs).

**Table 4.** PrEP Acceptability based on the Theoretical Framework of Acceptability.

| Construct | Question Asked | Response (Score) | Frequency | Proportions (%) |
| --- | --- | --- | --- | --- |
| Affective Attitude | How comfortable do you feel to use PrEP? | Comfortable (1) | 374 | 91.4 |
|  |  | Uncomfortable (0) | 35 | 8.6 |
| Burden | How much effort will it take to use PrEP? | Little/No effort (1) | 313 | 76.5 |
|  |  | A lot of effort (0) | 96 | 23.5 |
| Perceived Effectiveness | PrEP is likely to reduce my chance of getting infected with HIV | Agree (1) | 337 | 82.4 |
|  |  | Disagree (0) | 72 | 17.6 |
| Intervention coherence | It is clear to me how using PrEP reduces my chances of getting infected with HIV | Agree (1) | 380 | 92.9 |
|  |  | Disagree (0) | 29 | 7.1 |
| Self-Efficacy | How confident do you feel that you can do what is required to use PrEP | Confident (1) | 366 | 89.5 |
|  |  | Unconfident (0) | 43 | 10.5 |
| Opportunity Cost | Using PrEP will interfere with my other priorities | Agree (0) | 37 | 9.1 |
|  |  | Disagree (1) | 372 | 90.9 |
| PrEP Acceptability | Total of the 6 scores is 4 or more | Yes (Scores≥4) | 383 | 93.6 |
| | | No (Scores$<$4) | 26 | 6.4 |

### Factors associated with PrEP acceptability

Table 5 shows the factors associated with PrEP acceptability among the at-risk HIV negative young men in Kagwara fishing community. At a multivariable level, young men who perceived HIV risk had four times higher odds of accepting PrEP compared to their counterparts who never perceived HIV risk, aOR(95%CI) =4.23(1.05-17.04). Young men who knew their partners’ HIV status had lower odds of accepting PrEP compared to those who did not know their partners HIV status, aOR (95%CI) =0.25(0.25 (0.07-0.88). Those who felt embarrassed to ask for PrEP from the facility had lower odds of accepting PrEP, aOR(95%CI) =0.12(0.04-0.39), and young men who were concerned about stigma of using PrEP had lower odds of accepting PrEP, aOR (95%CI) = 0.13(0.04-0.41).

**Table 5.**
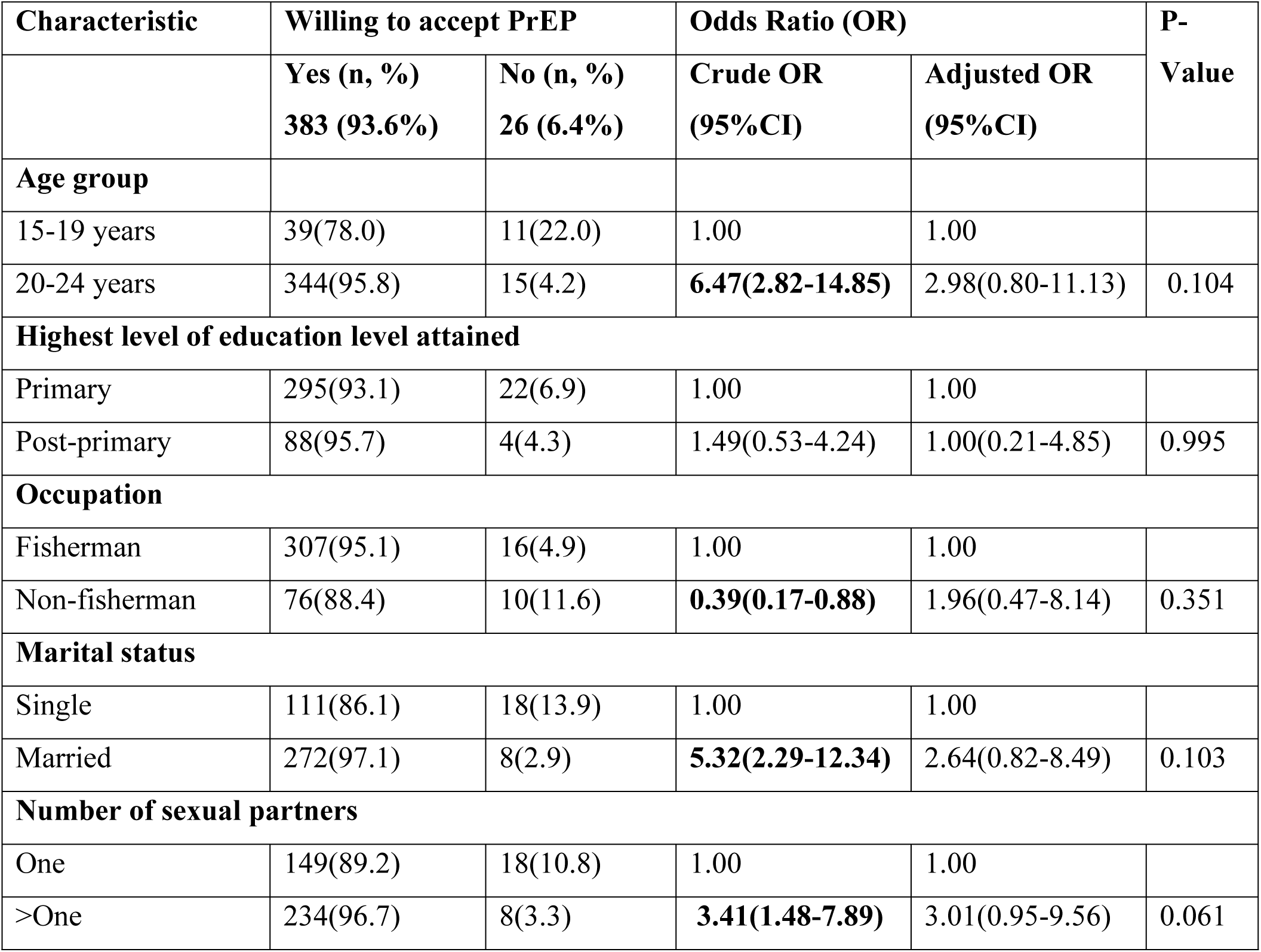
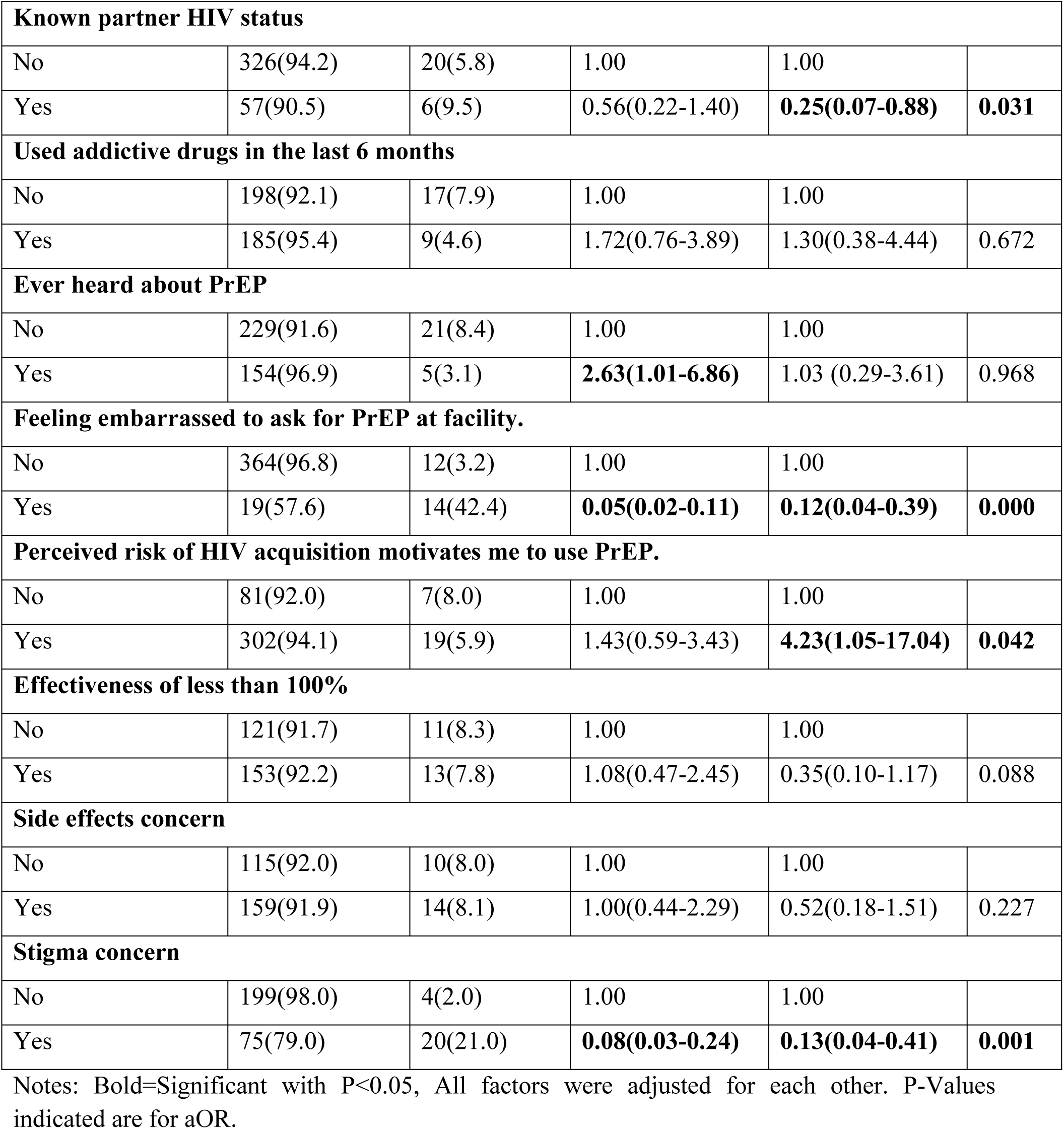
Factors associated with PrEP acceptability among at-risk, HIV-negative men in Kagwara fishing community.

| Characteristic | Willing to accept PrEP |  | Odds Ratio (OR) |  | P- |
| --- | --- | --- | --- | --- | --- |
|  | Yes (n, %)<br>383 (93.6%) | No (n, %)<br>26 (6.4%) | Crude OR<br>(95%CI) | Adjusted OR<br>(95%CI) | Value |
| Age group |  |  |  |  |  |
| 15-19 years | 39(78.0) | 11(22.0) | 1.00 | 1.00 |  |
| 20-24 years | 344(95.8) | 15(4.2) | 6.47(2.82-14.85) | 2.98(0.80-11.13) | 0.104 |
| Highest level of education level attained |  |  |  |  |  |
| Primary | 295(93.1) | 22(6.9) | 1.00 | 1.00 |  |
| Post-primary | 88(95.7) | 4(4.3) | 1.49(0.53-4.24) | 1.00(0.21-4.85) | 0.995 |
| Occupation |  |  |  |  |  |
| Fisherman | 307(95.1) | 16(4.9) | 1.00 | 1.00 |  |
| Non-fisherman | 76(88.4) | 10(11.6) | 0.39(0.17-0.88) | 1.96(0.47-8.14) | 0.351 |
| Marital status |  |  |  |  |  |
| Single | 111(86.1) | 18(13.9) | 1.00 | 1.00 |  |
| Married | 272(97.1) | 8(2.9) | 5.32(2.29-12.34) | 2.64(0.82-8.49) | 0.103 |
| Number of sexual partners |  |  |  |  |  |
| One | 149(89.2) | 18(10.8) | 1.00 | 1.00 |  |
| >One | 234(96.7) | 8(3.3) | 3.41(1.48-7.89) | 3.01(0.95-9.56) | 0.061 |
| **Known partner HIV status** |  |  |  |  |  |
| No | 326(94.2) | 20(5.8) | 1.00 | 1.00 |  |
| Yes | 57(90.5) | 6(9.5) | 0.56(0.22-1.40) | **0.25(0.07-0.88)** | **0.031** |
| **Used addictive drugs in the last 6 months** |  |  |  |  |  |
| No | 198(92.1) | 17(7.9) | 1.00 | 1.00 |  |
| Yes | 185(95.4) | 9(4.6) | 1.72(0.76-3.89) | 1.30(0.38-4.44) | 0.672 |
| **Ever heard about PrEP** |  |  |  |  |  |
| No | 229(91.6) | 21(8.4) | 1.00 | 1.00 |  |
| Yes | 154(96.9) | 5(3.1) | **2.63(1.01-6.86)** | 1.03 (0.29-3.61) | 0.968 |
| **Feeling embarrassed to ask for PrEP at facility.** |  |  |  |  |  |
| No | 364(96.8) | 12(3.2) | 1.00 | 1.00 |  |
| Yes | 19(57.6) | 14(42.4) | **0.05(0.02-0.11)** | **0.12(0.04-0.39)** | **0.000** |
| **Perceived risk of HIV acquisition motivates me to use PrEP.** |  |  |  |  |  |
| No | 81(92.0) | 7(8.0) | 1.00 | 1.00 |  |
| Yes | 302(94.1) | 19(5.9) | 1.43(0.59-3.43) | **4.23(1.05-17.04)** | **0.042** |
| **Effectiveness of less than 100%** |  |  |  |  |  |
| No | 121(91.7) | 11(8.3) | 1.00 | 1.00 |  |
| Yes | 153(92.2) | 13(7.8) | 1.08(0.47-2.45) | 0.35(0.10-1.17) | 0.088 |
| **Side effects concern** |  |  |  |  |  |
| No | 115(92.0) | 10(8.0) | 1.00 | 1.00 |  |
| Yes | 159(91.9) | 14(8.1) | 1.00(0.44-2.29) | 0.52(0.18-1.51) | 0.227 |
| **Stigma concern** |  |  |  |  |  |
| No | 199(98.0) | 4(2.0) | 1.00 | 1.00 |  |
| Yes | 75(79.0) | 20(21.0) | **0.08(0.03-0.24)** | **0.13(0.04-0.41)** | **0.001** |

Notes: Bold=Significant with $P < 0.05$ , All factors were adjusted for each other. P-Values indicated are for aOR.

## Discussion

This study assessed HIV PrEP acceptability and associated factors among the at-risk HIV negative young men in Kagwara fishing community-Serere district. Almost all (93.6%) of the respondents accepted PrEP for HIV prevention. This higher acceptability of PrEP shows a positive progress in HIV prevention efforts. Previous studies documented low PrEP acceptability among various populations. Among young men PrEP acceptability ranged from 45.45% to 76.80% in Sub-Saharan Africa (SSA) [26]. In Uganda, PrEP acceptability has been equally low in the earlier studies. For instance the study done in the fishing community of Lake Victoria found PrEP acceptability ranging from 14%-24% [27]. Subsequent studies have showed increasing PrEP acceptability among various populations. For example Bashir Ssuna et al reported 8 in 10 fisher folks in Kampala were willing to use PrEP [28]. Likewise, Susan S Witte reported 9 in 10 female sex workers were willing to initiate PrEP in Uganda [29]. This higher acceptance could have resulted from sustained behavior change messages targeting high risk communities such as fisher folks in the fishing communities and the adoption and promotion of PrEP as a key biomedical strategy for HIV prevention by ministry of health of Uganda [30]. The acceptability of PrEP by these groups therefore provides an opportunity for the ministry of health to expand access to PrEP services to reach the communities as one of a combination prevention strategy to reaching HIV epidemic control in Uganda. Ministry of health needs to use multiple approaches to provide PrEP such as peer-led models, drug distribution points, short message reminders for refills, pharmacies and retail drug shops

We found perceived risk of HIV infection as a positive factor associated with increased PrEP acceptability among the at-risk HIV negative young men in Kagwara fishing community. This implies that most of the young men engage in sexual relationships without knowing their partners’ HIV status. The perception here can be attributed to limited HIV testing in most of the communities in Uganda and young men practice unsafe sex which results in increased spread of HIV. This finding agrees with other studies that found a relationship between perceived risk of HIV and PrEP acceptability [31–33]. It is important to make PrEP services available along with HIV-tests and build capacity of the young men in performing HIV self-tests. In addition, various HIV testing modalities that are user friendly should be readily available like HIV self-testing at both facility and community.

Our study found knowing partners’ HIV status as one of the factors associated with reduced PrEP acceptability. This could be attributed to mutual trust developed by the partners. This finding is contrary with those of Ssuna et al. who did their study in Uganda and found knowledge of HIV status being associated with high PrEP use [34]. They reported that study participants who tested for HIV within the past 6 months were more likely to use PrEP. This requires packaging of HIV prevention messages in focused manner, educating young men about HIV window period and use of multiple HIV prevention methods like circumcision to cater for those that will not accept PrEP.

Our study also revealed that feeling embarrassed to ask for PrEP from the health facilities was another factor that reduced PrEP acceptability among the young men in Kagwara fishing community. This can be attributed to inferiority and high illiteracy levels common in the fishing communities in Uganda. This finding is in line with the study conducted among transgender women in U.S.A that identified being embarrassed with health care system as one of the factors affecting PrEP acceptability [35]. There is a need to train and orient health workers on customer care to handle such young men, introduce and sustain youth friendly health services at both health facilities and communities with focus in places where young men always have leisure activities.

In addition, our study found concern of stigma associated with PrEP use as another factor that reduced PrEP acceptability among the young men in Kagwara fishing community. Stigma has been associated with reduction of health care acceptability particularly in chronic illness like HIV. This finding has been also reported by study conducted among transgender women in U.S.A [36]. there is a need to break this barrier through continued sensitization of the young men to embrace the available HIV prevention methods.

## Study limitations and strengths

Our study had some limitations that included limited previous research studies on acceptability of PrEP by at-risk HIV-negative young men and this might have affected the theoretical foundation of our research, and this was managed by using studies on PrEP acceptability done in the other populations. We did not provide physical PrEP to study participants but only reported their presumed acceptability and in case they are to be given real PrEP pills, actual acceptability might reduce during implementation. The recruitment of only men affected overall acceptability level for the population and limited external validity and extrapolation of findings to the general population. Some participants might have had prior knowledge or education on PrEP and this might have insinuated some responses.

Despite these limitations, our study had strengths: This was among the few studies conducted among the at-risk HIV negative young men in the fishing community which is one of the unique populations prone to HIV, the knowledge gained from the present study may aid in lowering HIV infections among the at-risk HIV negative young men, findings can help to inform efforts to increase health promotion and continued sanitization to encourage young men to accept PrEP. Finally, findings in this study can serve as foundation for future researchers, policy makers and health professionals.

## Conclusion

Our study revealed that HIV PrEP is acceptable by at-risk HIV negative young men in Kagwara fishing community. The factors associated with HIV PrEP acceptability were perceived risk of HIV, knowing partner(s) HIV status, feeling embarrassed to ask for PrEP from health facilities and concern of stigma associated with PrEP use. These findings indicate the need to promote HIV PrEP among at-risk HIV negative young men in the fishing communities with intensified health promotion messages. There is a need to continue promoting use of multiple HIV prevention approaches and create awareness on the people eligible for PrEP.

It is important to identify barriers to PrEP acceptability among this population using a qualitative approach. This will help in designing person centered approach to PrEP delivery among young men in the fishing community.

## Data Availability

All relevant data are within the manuscript and its Supporting Information files.

## List of abbreviations

AGYW: Adolescent Girls and Young Women
AIDS: Acquired Immunodeficiency Syndrome
AVERT: Aid Virus Education and Research Trust
AYFRHS: Adolescent and Youth Friendly Reproductive Health Care Services
CDC: Centers for Disease Control and Prevention
DHIS2: District Health Information System Software version 2
HIV: Human Immunodeficiency Virus
HMIS: Health Management Information System
MSM: Men who have sex with Men
MSMW: Men who have Sex with Men and Women
NRTIs: Nucleotide Reverse Transcriptase Inhibitors
PrEP: Pre-exposure prophylaxis
PWID: People Who Inject Drugs
PWUD: People Who Use Drugs
STD: Sexually Transmitted Disease(s)
UN: United Nations
UNAIDS: United Nations Joint Program on HIV
UNFPA: United Nations Fund for Population Activities
UNICEF: United Nations International Children’s Emergency Fund
WHO: World Health Organization

## Declarations

### Ethical approval and consent to participate

Ethical approval to conduct the study was obtained from Busitema University Research and Ethics Committee (REC), Approval number **BUFHS-2023-61**. Administrative clearance was obtained from the district health office, Serere district and the local government representative office in whose jurisdiction Kagwara landing site was situated. Given that the study was assessing behavior related to HIV prevention among high-risk HIV-negative persons, this certainly needed to uphold several ethics when conducting the study. Each participant at his own discretion provided an informed consent (written) as was indicated on the consent form, while parental consent was obtained for all the minors prior to their participation in the study. Confidentiality was maintained by ensuring that individual patient-level data obtained was de-identified, encrypted, and passworded to ensure access by only an authorized team of investigators. All interviews were conducted in a place where the interviewer was sure that there were no third parties listening to the interview. All the young persons sampled were informed that their participation in the study was voluntary, with the possibility of withdrawing from the study without any negative ramifications.

### Consent for publication

Not applicable

### Availability of data and materials

All relevant data are within the manuscript and its Supporting Information files.

## Authors’ contributions

AO, PO and JKBM conceptualized and developed the proposal. AO and JKBM supervised data collection. RO and FO conducted data curation and formal analysis. RO and AO drafted the manuscript. PO and JKBM reviewed the final manuscript. All authors read and approved the final manuscript.

## Acknowledgements

The authors would like to acknowledge the contribution of all the staff of the Department of Public and Community Health, Faculty of Health Sciences, Busitema University for their support during the design and implementation of the study. We would also like to appreciate the technical support from Dr.Ogwal Daniel, the district health officer Serere district who provided administrative clearance and oversaw data collection process in the landing site.

